# Fractional SIR Epidemiological Models

**DOI:** 10.1101/2020.04.28.20083865

**Authors:** Amirhossein Taghvaei, Tryphon T. Georgiou, Larry Norton, Allen Tannenbaum

## Abstract

The purpose of this work is to make a case for epidemiological models with fractional exponent in the contribution of sub-populations to the transmission rate. More specifically, we question the standard assumption in the literature on epidemiological models, where the transmission rate dictating propagation of infections is taken to be proportional to the product between the infected and susceptible sub-populations; a model that relies on strong mixing between the two groups and widespread contact between members of the groups. We content, that contact between infected and susceptible individuals, especially during the early phases of an epidemic, takes place over a (possibly diffused) boundary between the respective sub-populations. As a result, the rate of transmission depends on the product of fractional powers instead. The intuition relies on the fact that infection grows in geographically concentrated cells, in contrast to the standard product model that relies on complete mixing of the susceptible to infected sub-populations. We validate the hypothesis of fractional exponents i) by numerical simulation for disease propagation in graphs imposing a local structure to allowed disease transmissions and ii) by fitting the model to a COVID-19 data set provided by John Hopkins University (JHUCSSE) for the period Jan-31-20 to Mar-24-20, for the countries of Italy, Germany, Iran, and France.

## I. Introduction

The classical SIR (Susceptible, Infectious, Recovered) model of infectious disease dynamics, and all subsequent multi-compartmental derivative models, are based on a model for the transmission rate that is taken universally in the form

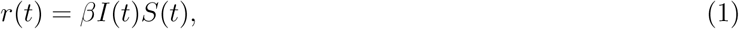

where *I*(*t*), *S*(*t*) represent the size of infected and susceptible sub-populations. The proportionality factor *β* is typically determined on a case-by-case basis. Thus, if *R*(*t*) represents the size of the recovered population, assuming that all individuals undergo full recovery and thereby the total population *S*(*t*) *+ I*(*t*) *+ R*(*t*) remains constant, the most basic model for transmissions is in the form of the following system of equations, known as ***SIR model***,

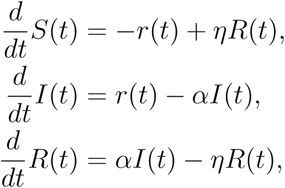

where *α* is the recovery rate (with time constant of recovery *τ :=* 1/*α*) and *η* a parameter regulating the rate at which immunity is lost over time. See [1], [11] for all the details about the SIR model together with an extensive list of references. Multi-compartmental models that include infected but asymptomatic individuals, deceased, etc. as well as a flux from the recovered to the susceptible sub-population, as immunity wears out, have also been considered. However, throughout, the basic feedback that drives the infection, *r*(*t*), is invariably as in (1).

In departure from this well-studied SIR paradigm, we propose a *fractional SIR* (***fSIR***) *model* with rate

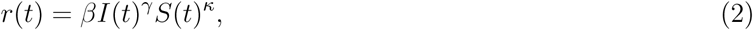

where one or, possibly, both sub-populations are scaled by exponents that are typically less than 1. The justification for such a model stems from the fact that, at least during the initial phase of an epidemic, infection propagates outwards from infected cells to the general population. In such a scenario, where for instance *S*(*t*) ≫ *I*(*t*) (much greater), the boundary of infected cells which would roughly account for most new infections, scales as a fractional power *γ <* 1 of the area of the cells, hence *I*(*t*)*^γ^*. In actuality, due to the diffusive nature of infection-propagation amongst the general population, the exponent is expected to be larger than 1/2, as it would be in the continuous limit when the boundary is a smooth curve. Moreover, at least in the early phases of an epidemic, the exponent of *S*(*t*), which is significantly larger than *I*(*t*) may turn out to be negligible.

The idea of using fractional exponents in growth models has been motivated from Norton-Simons-Massagué (NSM) model, a growth model of the form

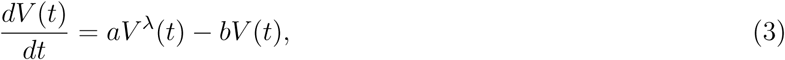

with origins in the 1950s [3]. This type of model was designed to describe the growth of biological organisms employing certain energy principles. In the model, the parameters *a* and *b* quantify anabolism (growth) and catabolism (death), respectively. Equation (3) may be interpreted as asserting that the net growth rate of an organism results from the balance of synthetic and degradative mechanisms. While the rate of the former process follows a law of allometry (i.e., the rate is proportional to the volume *V*(*t*) via a power function), the rate of the latter process scales linearly with *V*(*t*). It is important to note that the two special cases of (3), (i) power law *b* = 0, and (ii) second-type growth *λ* = 2/3 have already been successfully applied to describe tumor growth [15], [6]. The general case, 0 < *λ* < 1, was introduced in [14] to explain the self-seeding hypothesis. Moreover, an important geometrical interpretation was provided in [13], [2]. In these works, the authors relate the exponent *λ* = *d*/3 to the fractional Hausdorff dimension of the proliferative tissue, where *d* denotes the fractal dimension of the tissue. Moreover, the model (3) has been derived mechanistically by linking tumor growth to metabolic rate and vascularization [8].

In a similar spirit to the Norton-Simons-Massagué (NSM) model, herein, we recognize the geometric constraints imposed on disease propagation by the locality of transmissions around infectious cells. To this end, we seek to explain the origin of fractional power in (2) by i) numerical experiments, and ii) fitting such models to data sets.

Specifically, with respect to i), we postulate a discrete model where infection propagates over nodes of a network. The network, representing individuals, is not planar (in general), yet it is immersed in 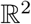. While (physically) neighboring nodes may be densely connected, precluding the graph from being planar, the likelihood of being connected as well as infecting each other depends on their physical distance. It is observed that such models lead to fractional exponents in the transmission rate, in agreement with (2). With regard to ii), we have also numerically studied data on recent COVID-19 epidemic that is available at [5]. For this dataset, as we note in the results, the exponent *γ* in (2) ranges from about 0.6 to 0.8, which is similar to the empirically determined exponent of the NSM model.

## II. Models of discrete transmissions

To provide insight and justification for our hypothesis on the validity of equation (2), we develop a discrete model for direct transmissions between individuals consisting of nodes (individuals) on a graph that captures contacts between them. In the present work, the graph is fixed, while in future work we plan to explore the possibility of time-varying links between nodes as well as modeling control actions, such as social-distancing, so as to study the effects of such mediation-protocols.

### A. Model: probabilistic SIR on a graph

Consider a simple graph of size *n* with adjacency matrix *A*. The graph is used to model the spread of infection over a network of nodes representing individual people. Every node can be in one the three states {*S, I, R}*. We use *x_i_*(*t*) ∊ {*S, I, R}* to represent the state of node *i* at time *t*. Here, *x_i_*(*t*) evolves, as a Markov chain on 3*^n^* states, according to the following transition probabilities at time *t* dictating transition at the node level,

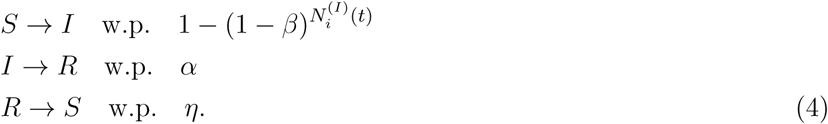

Here, *β* is the infection rate, *α* is the recovery rate, and *η* is the susceptibility rate (quantifying loss of immunity over time). The notation 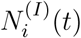 stands for the number of the neighbors of node *i* that are infected at time *t*, i.e.

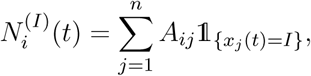

where 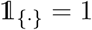 when {·} holds and is 0 otherwise.

With regard to the structure of the graph, specifying contacts between individuals, we describe results considering the following options:

*1) Two-dimensional grid-graph:* We work out two rudimentary models where individuals (nodes) are placed on a 2-dimensional grid (vertex set)

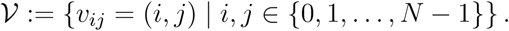

We carry out experiments for two cases, where the edge set is defined by

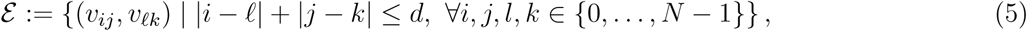

with *d ∊* {1, 2}. Thus, when *d* = 1, each node is connected to *k* = 4 nearest neighbors, while in the case where *d =* 2 each node is connected to *k =* 8 nearest neighbors. The graph structure for the case *d* = 1 is depicted in Figure 1a. In either case, the infectious model is simulated and the results discussed in the section on experiments.

**Fig. 1:**
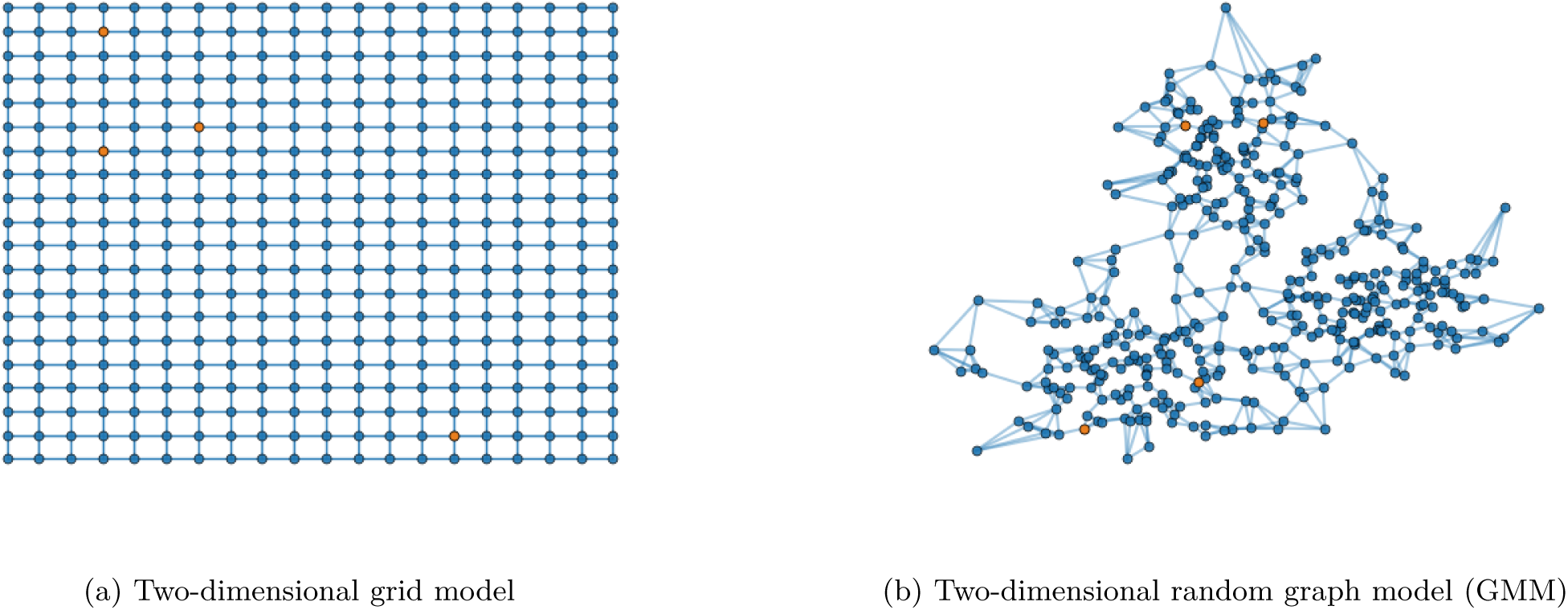
Structure of the two graph models considered in this paper. Nodes are connected to their 4 nearest neighbors. The initially infected people are marked in orange.

*2) Two-dimensional random graph:* We postulate a distribution of nodes on 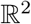 according to a Gaussian-mixture model (Figure 1b). Each node is connected to its 4 nearest neighbor. Analogous conclusions are drawn and discussed in the section on experiments as well.

We experiment with a randomized initialization, where a number of small initial cells of infected individuals are sprinkled randomly with probability *p*_0_ inside the general population. A localized initialization, where a specified initial collection of neighboring nodes are infected and the contagion begins from these nodes, gives similar results.

## III. Experiments

### A. Two-dimensional grid-graph

Simulation results of infection spread on the two-dimensional grid-graph with size 100 × 100, for *d* ∊ {1, 2} (and hence, each node connected to the *k* ∊ {4,8} nearest nodes), were carried out for the model (4) and are presented in Figure 2–3 and Figure 4–5, respectively. The documented experiments were carried out for *η =* 0.01, initial infection probability *p*_0_ *=* 10^−3^ and combination of parameters for *α =* {0.05, 0.1} and *β* ∊ {0.2, 0.3}.

**Fig. 2:**
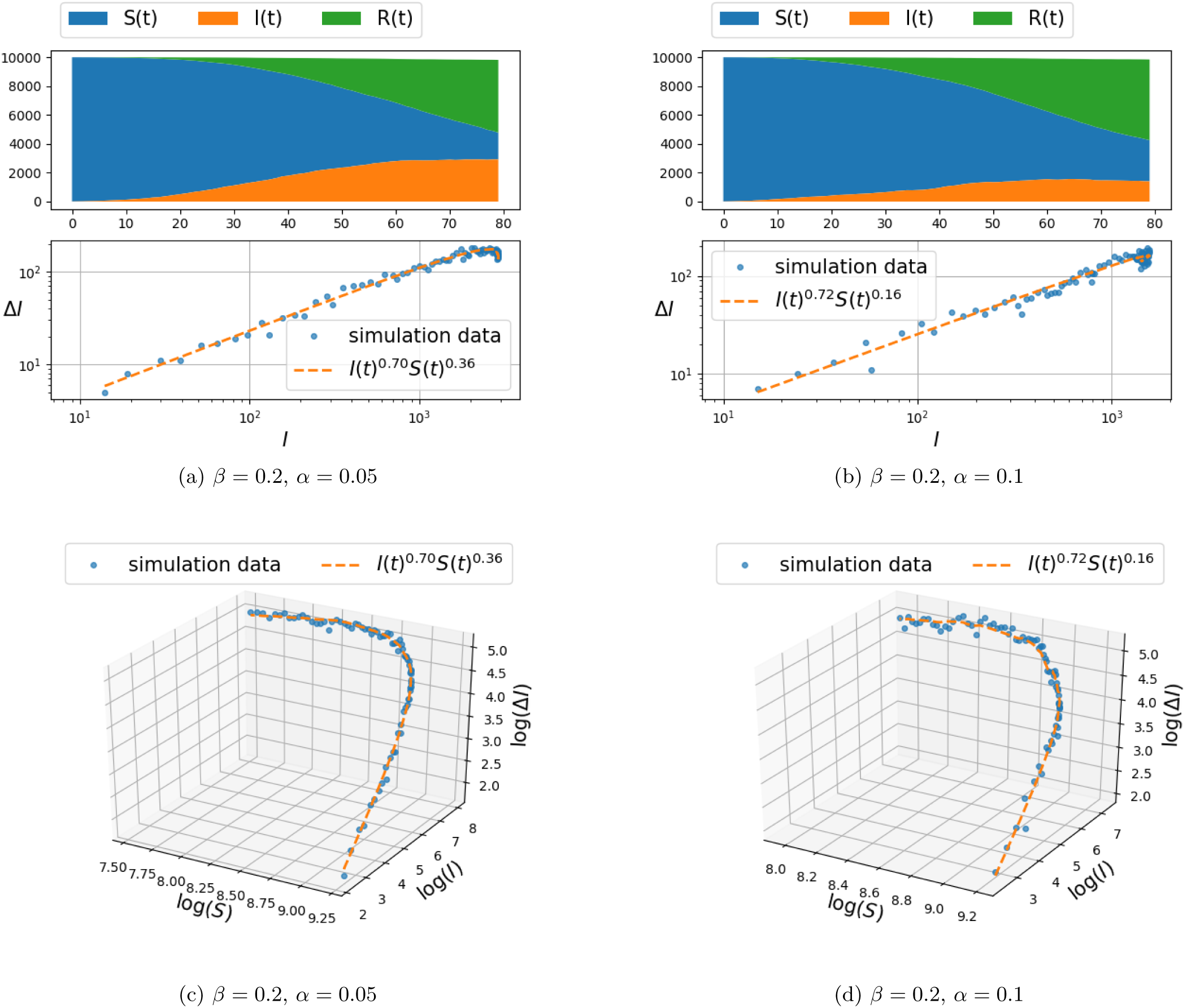
Spread of infection on a two dimensional grid, with connection to *k* = 4 nearest neighbors (i.e., *d* = 1 in Eq. (5)) and *β* = 0.2.

**Fig. 3:**
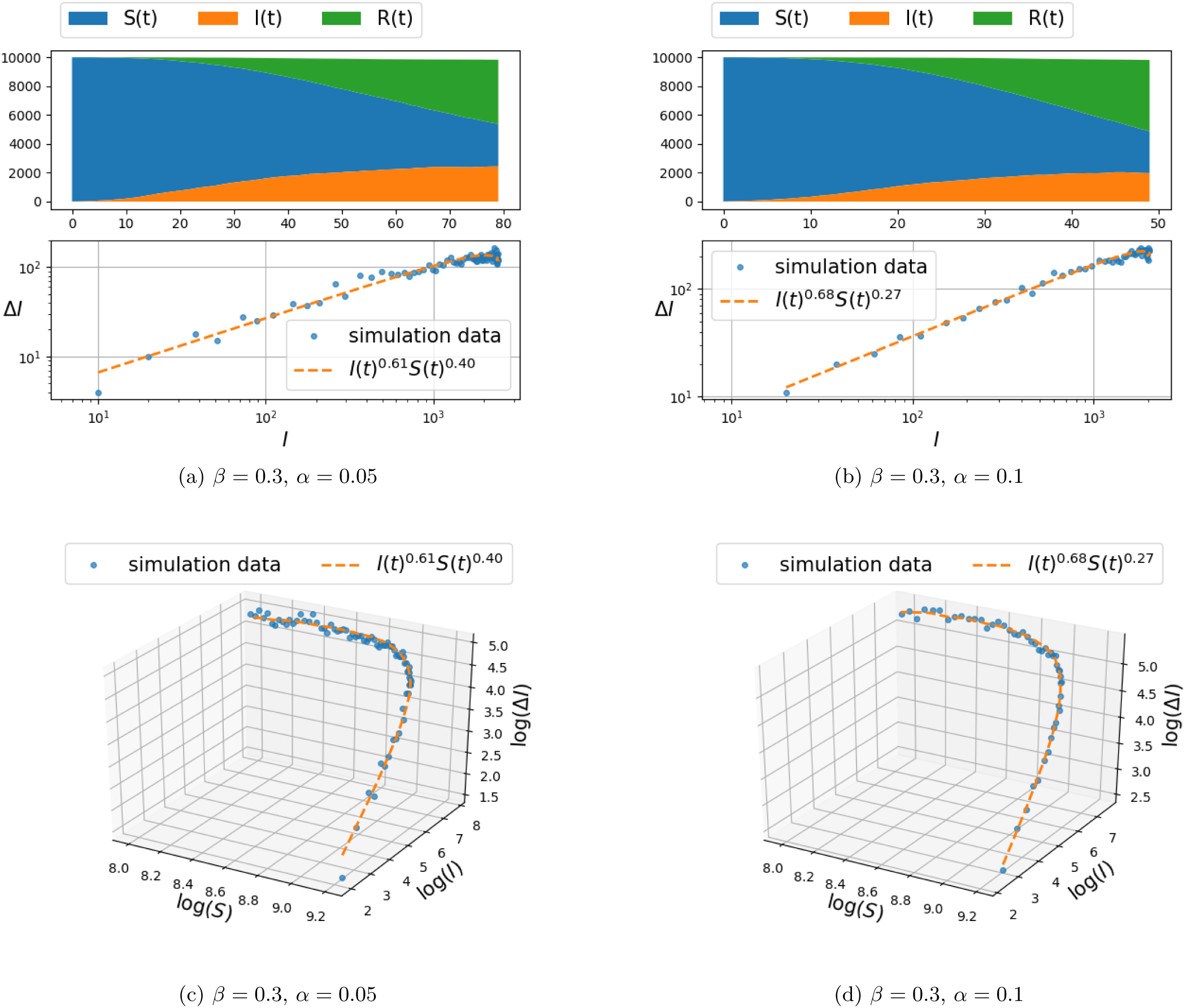
Spread of infection on a two dimensional grid, with connection to *k* = 4 nearest neighbors (i.e., *d* = 1 in Eq. (5)) and *β* = 0.3.

**Fig. 4:**
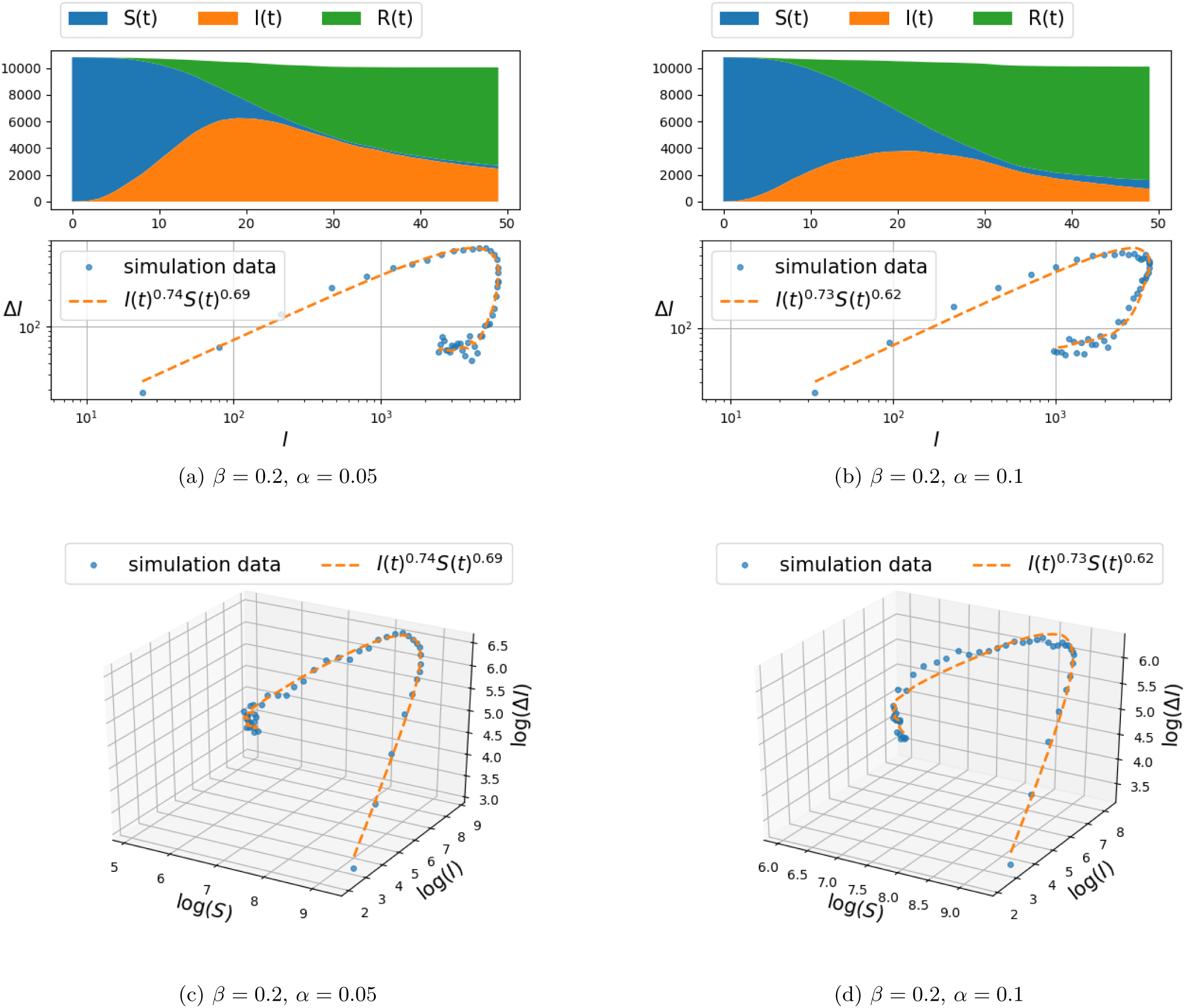
Spread of infection on a two dimensional grid, with connection to *k* = 8 nearest neighbors (i.e., *d* = 2 in Eq. (5)) and *β* = 0.2.

**Fig. 5:**
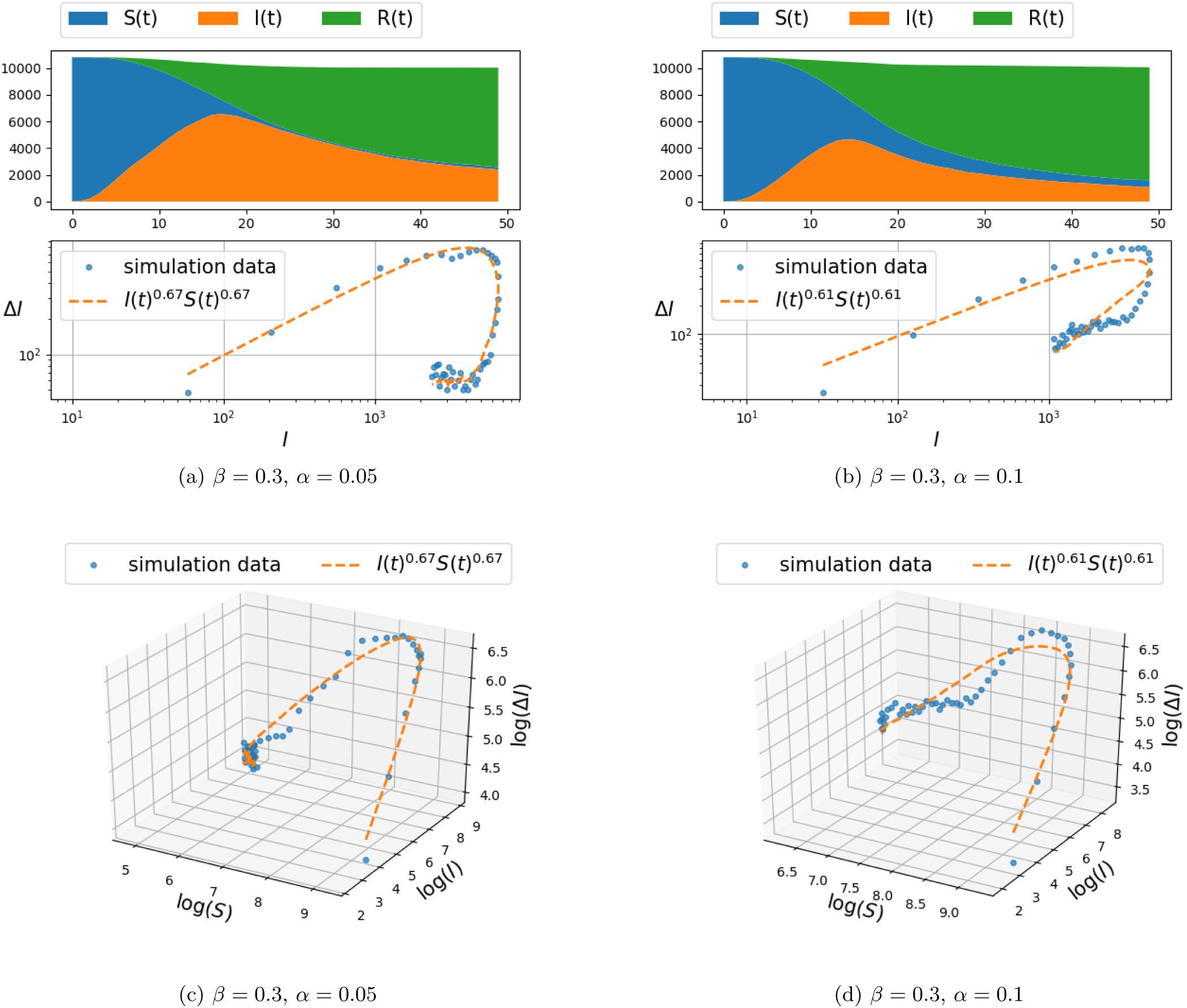
Spread of infection on a two dimensional grid, with connection to *k* = 8 nearest neighbors (i.e., *d* = 2 in Eq. (5)) and *β* = 0.3.

The populations of susceptible, infected, and recovered people are calculated as follows:

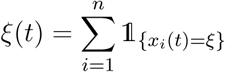

for each ξ *∊ {S, I, R}*. The top panel in each figure depicts the number of susceptible, infected, and recovered individuals as a function of time. The number of newly infected population at time t, denoted by Δ*I*(*t*), vs. the number of infected people *I*(*t*) is shown in the second panel (marked by dots). To be precise, Δ*I*(*t*) is defined according to

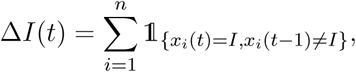

and thereby, Δ*I*(*t*) only includes the newly infected people.

In order to capture the relationship between Δ*I*(*t*) and *I*(*t*) and *S*(*t*), a parametric curve of the form Δ*I*(*t*) *= cI*(*t*)*^γ^S*(*t*)*^κ^*, is fitted to the data-points obtained from the simulation. The constant c, and the exponents *γ* and *κ* are obtained from a least-squares method applied to linear relation between respective logarithms^1^,

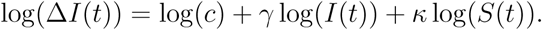

The two-dimensional projection of the curve on the (log(Δ*I*), log(*I*)) plane is depicted in the second layer of panels and compared to the distribution of (Δ*I*(*t*)*,I*(*t*)) point set. The third layer of panels compares the fit of curve to the (Δ*I*(*t*)*,I*(*t*)*,S*(*t*)) data set in a three-dimentional plot in logarithmic scales.

### B. Two-dimensional random graph

Simulation result on a two-dimensional random graph model were carried out and documented in Figure 6. The spatial distribution of 10^4^ node-coordinates ((*x*, *y*)-coordinates, cf. Figure 1b) has been selected from the mixture of three Gaussians distributions^2^ on the node-coordinate plane,

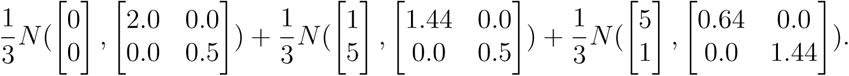

**Fig. 6:**
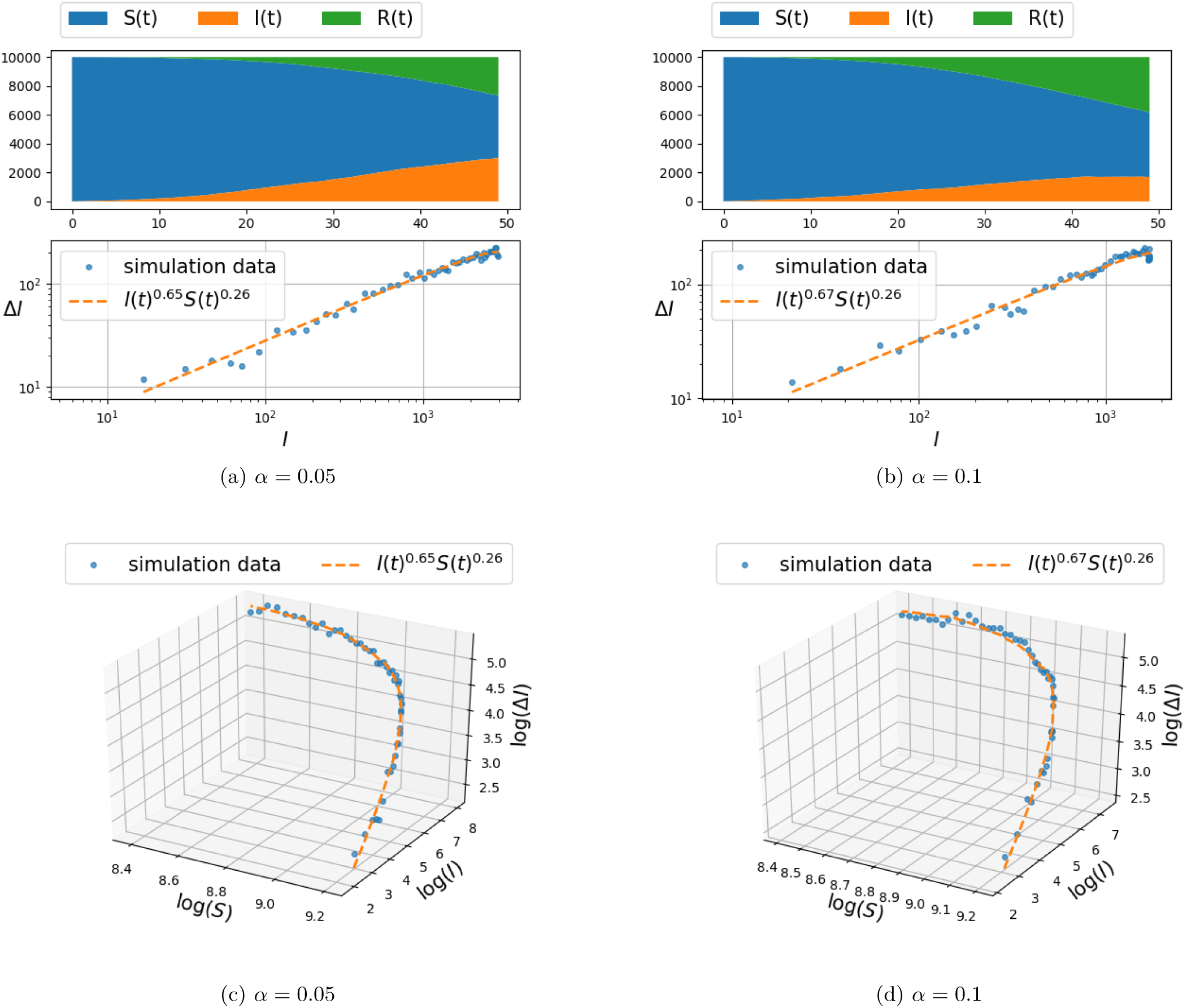
Spread of infection on a mixture of Gaussian random graph model for *β* = 0.3.

Initially, for the experiment documented in Figure 6, the nodes are connected to their 4 nearest neighbors. The infection spread model is once again the one specified in (4). The parameters selected for the results in Figure 6 are: (*β=* 0.3 and *η =* 0–01, while we compare the effect of *α* ∊ {0.05, 0.1}. As before, the first layer of panels depicts the number of susceptible, infected, and recovered individuals as a function of time. The two-dimensional projection of the curve on the (log(Δ*I*), log(*I*)) plane is depicted in the second layer of panels and compared to the distribution of (Δ*I*(*t*)*,I*(*t*)) point set, while the third layer of panels compares the fit of curve to the (Δ*I*(*t*)*,I*(*t*)*,S*(*t*)) data set in a three-dimensional plot once again in logarithmic scales.

Two additional plots that are based on the same random spatial distribution of nodes are shown in Figure 7.

**Fig. 7:**
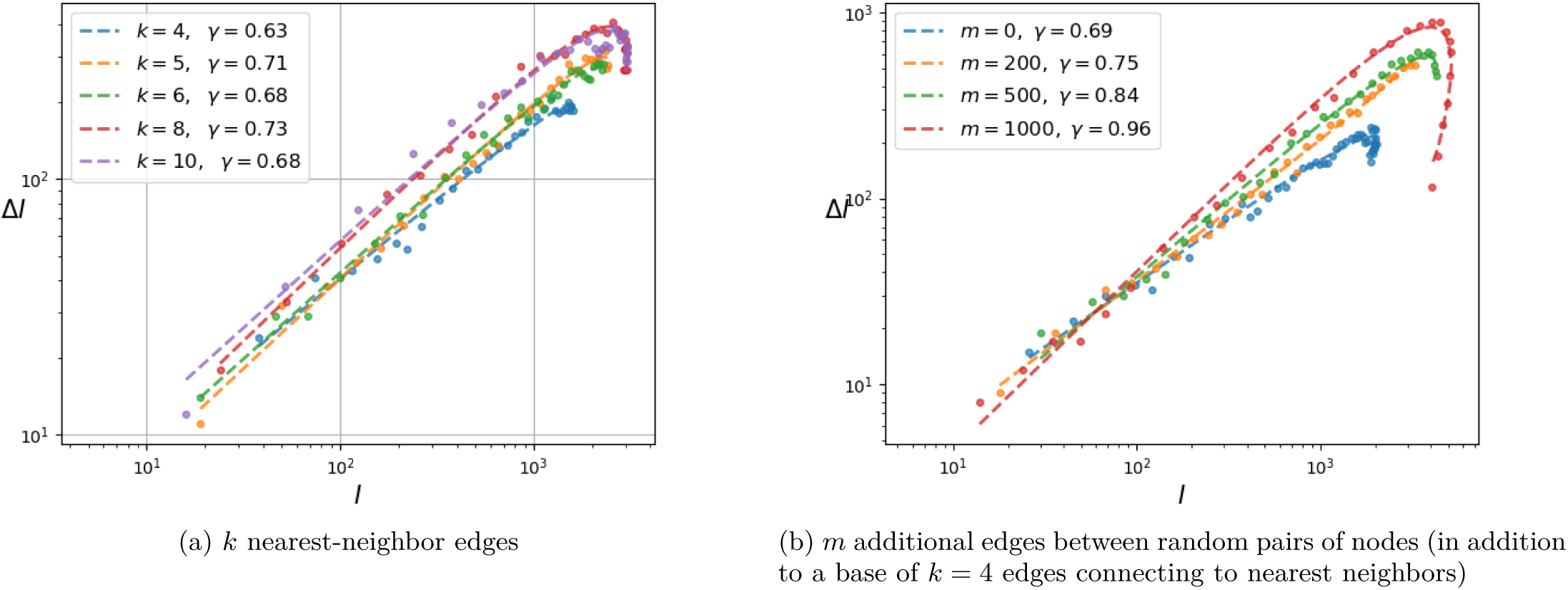
Dependence of *γ* on the graph connectivity.

On the left panel, a range of values for the parameter *k* ∊ {4, 5, 68, 10}, that represents the number of nearest neighbors connected to each node, is explored in its impact on the exponent *γ* of the (Δ*I, I*) relation. It is observed that *k* has only a very small effect, as *γ* ranges between 0.63 – 0.73.

On the right panel, a different experiment is carried out, introducing additional edges randomly that connect (even remote) pairs of nodes. Here *m* ∊ {0, 200, 500, 1000} designates the number of random edges in this graph of 10^4^ nodes. The random introduction of edges, beyond the original *k =* 4 nearest neighbor connections, decreases the diameter of the graph (longest distance between any two nodes) and increases rapidly the exponent *γ*.

### C. COVID-19 data-set

We utilized data provided by the Johns Hopkins University Center for Systems Science and Engineering [5]. We study the relationship between the number of newly infected Δ*I*(*t*) and the total recorded number of infected *I*(*t*) individuals for the duration 01/31 to 03/24, in Italy, Germany, Iran, and France. The number of newly infected individuals is approximated by subtracting the number of infected people at time *t* from the number of infected people at time *t* − 1, as information on those recovering is not available. We fitted the curve Δ*I*(*t*) *= cI*(*t*)*^γ^* to the data; due to the fact that *S*(*t*) ≫ *I*(*t*) during these initial stages of infection spread *S*(*t*) is treated as constant. The result for four different countries is depicted in Figure 8.

**Fig. 8:**
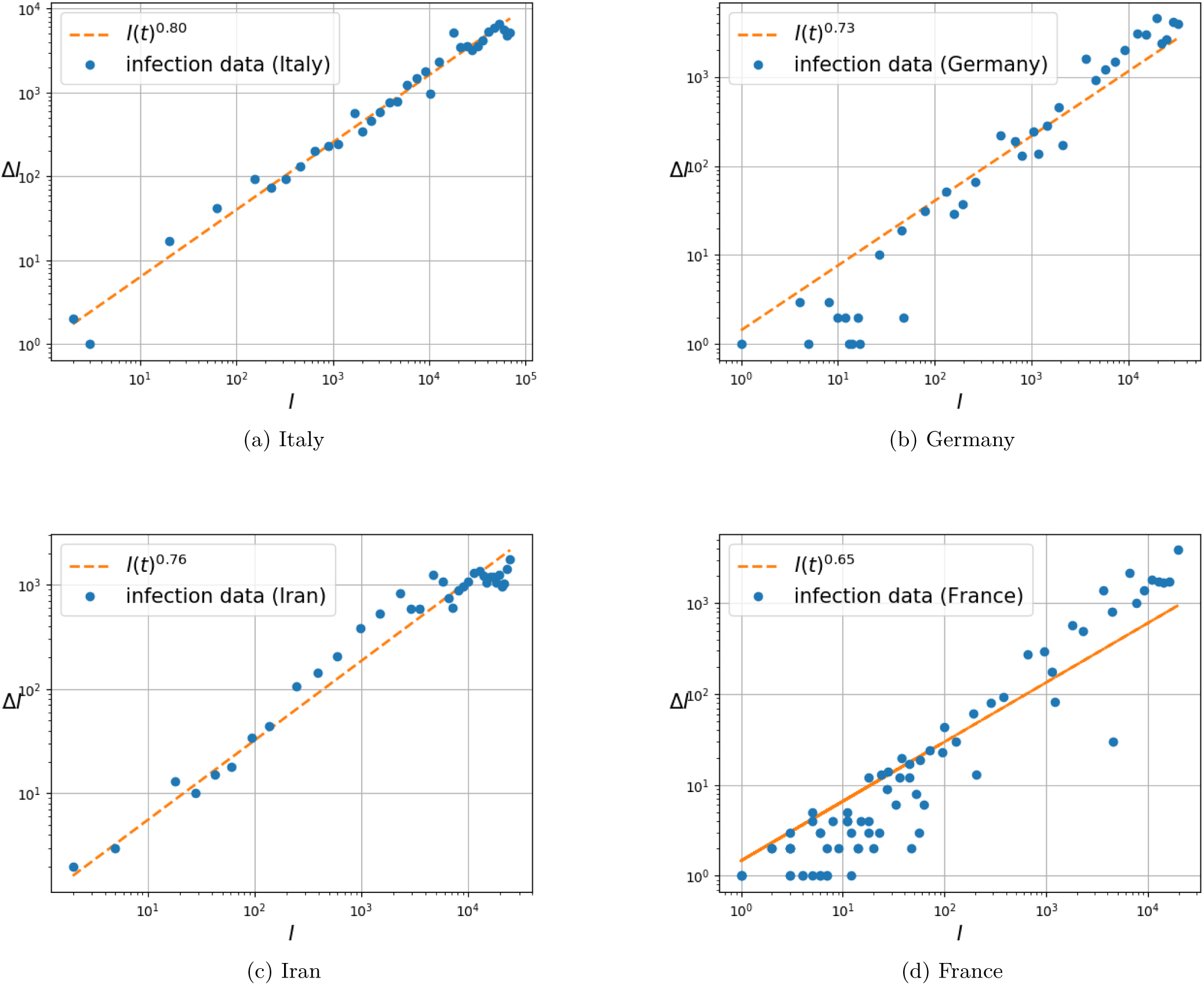
Relationship between infection growth Δ*I*(*t*) and the number of infected individuals *I*(*t*) with COVID-19 in four different countries.

## IV. Discussion

The recent onset of the COVID-19 pandemic has underscored the urgency of accurate models for the spread of infectious diseases. These help guide the allocation of resources, intervention, and mediation strategies, and may help quantify the impact of lifestyle changes on the progression of the epidemic and the threshold for herd immunity. In particular, one immediate practical question in the current COVID-19 pandemic is to decide when an intervention—such as the need to reinstitute social distancing (that had been relaxed)—is appropriate. Thus, while efforts to develop accurate models go back almost a century [10] (see also [4], [9]), the subject is especially urgent today.

The main thesis of this work is that models of epidemics, especially during the early phases, incorrectly assume that the contagion depends on the product of infected and susceptible populations. Contagion takes place at the boundary of infected cells and as a result it is the topology of the distribution of infected cells that dictate the spread. An analogous situation takes place in tumor growth, where models suggest (see [3], [12], [14]) fractional exponent for the contribution of tumor volume, as this more accurately captures the size of the boundary that affects growth. Thus, based on an analogous rationale, we propose a fractional-power alternative, fSIR, to the standard SIR model of disease dynamics. The value of exponents depend on a number of factors including the nature of the boundary between infected cells and the general susceptible population. Specifically, the exponent relates to the level mixing between infected and healthy individuals at interface between the two sub-populations, and may be quantified by the diameter of the graph that represents contacts between individuals.

Our thesis is supported by simulation results as well as by fitting this fSIR model to recent COVID-19 datasets. Specifically, the two-dimensional discrete probabilistic SIR models in Figures 2–3 (with *k* = 4 nearest-neighbor connection) and in Figures 4–5 (with *k =* 8 nearest-neighbor connection), that simulate disease propagation on a discrete domain of nodes (representing individuals in contact with one another), suggest exponents 7 in the range between 0.61 − 0.74 for the contribution of *I*(*t*) on the infection rate. Similar results are observed in Figure 6 for a two-dimensional random distribution of nodes (vertex set) with four nearest-neighbor contacts (edge set). Here, the exponent of the *I*(*t*) contribution to the infection rate lies in a similar range ({0.65, 0.67} for the conditions displayed). Two additional plots in Figure 7 highlight the weak dependence of *γ* on the number of short-range contacts (nearest neighbors) and the strong dependence on even a few long-range contacts amongst the general population.

The fit of the COVID-19 data-set [5] gives exponents for the contribution of *I*(*t*) on the infection rate in the range of 0.65 − 0.8. In this data-set, the value of *S*(*t*) (that includes the remaining of a rather large total population) varies insignificantly over time, and hence may be treated as constant. Several limitations of our experiment are noted. Firstly, the value of *I*(*t*) is only an estimated value since recording of *all* infected individuals is not guaranteed. Secondly, the value of Δ*I*(*t*) is estimated as being the difference *I*(*t*) − *I*(*t −* 1); we cannot take into account individuals who may have recovered. However, it is deemed that the uncertainty in the actual value of *R*(*t*) that quantifies the recovered sub-population is not significant; this can be argued based on the basis that the COVID-19 recovery period is of the order of weeks.

It is anticipated that, in a similar manner as in the Norton-Simon hypothesis on cancer treatment [7], the rate of regression under effective intervention is proportional to the expected rate of growth of a population of that size, and that the most efficient intervention would be dose dense, either continuous if possible or as frequently applied as possible. This is important since should relaxation of social distancing be allowed to proceed too long before re-institution of same, this would be predicted to be disadvantageous.

The authors believe that it is imperative that a deeper and more extensive study is carried out, whereupon the values of *I*(*t*), Δ*I*(*t*), *R*(*t*) are estimated from more extensive datasets. The effect of mediation efforts, such as social distancing, should be recorded as well and taken into account by differentiating data for the periods before and after such mediation protocols take effect. It is the authors’ hope that questions raised in this work, as to the validity of the basic assumption in SIR models, lead to more reliable and robust ways to estimate the progression of epidemics as well as the progression of the current COVID-19.

## Data Availability

Data used are publically available.

https://github.com/CSSEGISandData/COVID-19

## Acknowledgments

This research was supported by AFOSR Grants FA9550-20-1-0029, FA9550-17-1-0435, NSF Grants 1807664, 1839441, National Institute of Aging Grant R01-AG048769, MSK Cancer Center Support Grant/Core Grant (P30 CA008748), and a grant from Breast Cancer Research Foundation BCRF-17-193.

## Footnotes

1 The zero entries for Δ*I*(*t*)*,I*(*t*)*,S*(*t*) are ignored.

2 *N*(*v, R*) denotes a Gaussian distribution with mean *v* and covariance *R*.

## References

[1] F. Bauer, C. Castillo-Chavez, and Z. Feng. Mathematical Models in Epidemiology, volume 69. Springer, 2019.

[2] S. Benzekry, C. Lamont, A. Beheshti, A. Tracz, J M.L. Ebos, L. Hlatky, and P. Hahnfeldt. Classical mathematical models for description and prediction of experimental tumor growth. PLoS Computational Biology, 10(8):e1003800, 2014.

[3] L. Von Bertalanffy. Quantitative laws in metabolism and growth. The Quarterly Review of Biology, 32(3):217–231, 1957.

[4] Ottar N Bjørnstad, Katriona Shea, Martin Krzywinski, and Naomi Altman. Modeling infectious epidemics. Nature methods, 2020.

[5] Johns Hopkins University Center for Systems Science and Engineering. 2019 Novel Coronavirus COVID-19 (2019-nCoV) Data Repository. https://github.com/CSSEGISandData/COVID-19, 2020. [Online; accessed 25-April-2020].

[6] P. Gerlee. The model of muddle: in search of tumor growth laws. Cancer Research, 73:2407–2411, 2013.

[7] Richard Gray, Rosie Bradley, Jeremy Braybrooke, Zulian Liu, Richard Peto, Lucy Davies, David Dodwell, Paul McGale, Hongchao Pan, Carolyn Taylor, et al. Increasing the dose intensity of chemotherapy by more frequent administration or sequential scheduling: a patient-level meta-analysis of 37 298 women with early breast cancer in 26 randomised trials. The Lancet, 393(10179):1440–1452, 2019.

[8] A. B. Herman, V. M. Savage, and G. B. West. A quantitative theory of solid tumor growth, metabolic rate and vascularization. PLoS ONE, 6(9):e22973, 2011.

[9] Herbert W Hethcote. The mathematics of infectious diseases. SIAM review, 42(4):599–653, 2000.

[10] William Ogilvy Kermack and Anderson G McKendrick. A contribution to the mathematical theory of epidemics. Proceedings of the royal society of london. Series A, Containing papers of a mathematical and physical character, 115(772):700–721, 1927.

[11] Maia Martcheva. An Introduction to Mathematical Epidemiology, volume 61. Springer, 2015.

[12] L. Norton. A gompertzian model of human breast cancer growth. Cancer Research, 48:7067–7071, 1988.

[13] L. Norton. Conceptual and practical implications of breast tissue geometry: Toward a more effective, less toxic therapy. The Oncologist, 10:370–381, 2005.

[14] L. Norton and J. Massagué. Is cancer a disease of self-seeding? Nature Medicine, 12(8):875–878, 2006.

[15] V. G. Vaidya and F. J. Alexandro. Evaluation of some mathematical models for tumor growth. International Journal of Bio-Medical Computing, 13:19–35, 1982.

